# Lower-limb Monitoring with IMUs in Sport: A Systematic Review

**DOI:** 10.1101/2025.07.11.25331348

**Authors:** AJ Lamb, Timothy J. Suchomel, Jennifer Strickler

**Affiliations:** Concordia University Chicago; Chicago Bears Football Club; University of Pittsburgh; Oklahoma City Thunder

**Keywords:** External load, inertial measurement units, athlete monitoring, wearables

## Abstract

The advancement of wearable technology has enhanced athlete monitoring across various sports and competitive levels. Inertial measurement units (IMUs) enable the quantification of external load at different anatomical locations, providing ecologically valid data in real-world sporting environments. This systematic review examines the prevalence, application, and methodological considerations of lower limb-worn IMUs in competitive sports outside laboratory settings. A comprehensive search across four databases identified 71 relevant studies categorized by publication information, participant demographics, device specifications, and task characteristics. Findings indicate a substantial increase in the use of lower limb IMUs over the past decade, with a wide range of spatiotemporal parameters analyzed across diverse athletic populations. Despite this growth, gaps remain in device specifications, longitudinal studies, and standardization of monitoring protocols. These results highlight the potential of IMUs as a noninvasive tool for spatiotemporal parameters and sport-specific movement patterns, offering valuable insights to refine training prescriptions and optimize athlete performance.

## 1. Introduction

Recent technological advances have led to the development of a wide range of wearable devices capable of quantifying internal and external load in applied athletic settings (1). These devices have enabled the implementation of athlete monitoring programs across team and individual sports, spanning all competitive levels, to inform training prescriptions and enhance performance outcomes [2, 3]. Among these wearables, inertial measurement units (IMUs) are commonly utilized to measure acceleration (e.g. accelerometers), rotation (e.g. gyroscopes), and orientation (e.g. magnetometer), typically sampling anywhere from 50-1600 Hz (2–4). A key advantage of IMUs is their ability to function independently of global navigation satellite systems (GNSS) or local positioning systems (LPS), allowing for the development of compact, lightweight sensors – measuring only 44 mm in length and weighing less than ten grams (2). Due to their lightweight, unobtrusive design, these sensors have been deployed at various anatomical sites to better understand sport-specific movement at the lower limb, including the ankle/tibia (5–10) and foot (11, 12).

Traditionally, whole-body external load has been quantified using wearable IMUs or positioning technology located at the thoracic spine. However, emerging evidence that external load is highly specific to the placement of the wearable, with thoracic-worn units often misrepresenting loads experienced at the lower limb (13). Consequently, this discrepancy may lead to an underestimation of the mechanical stress endured by distal tissues, which could contribute to suboptimal training prescriptions, impaired performance, and an increased risk of injury (14–16). Given these limitations, practitioners should critically evaluate the interpretation of whole-body external load metrics derived from positioning technologies or IMUs, as these measures may not be directly applicable to other anatomical regions. By reconsidering conventional approaches to load monitoring, practitioners may develop more targeted strategies for periodization or training, ultimately optimizing athlete performance.

Despite the increasing use of IMUs in sports science, research investigating their application in quantifying lower-limb-specific external load outside controlled laboratory environments remains limited (17). Existing studies conducted in field settings predominantly focus on distance-running tasks and recreationally active populations (13, 18, 19). As a result, there is a notable gap in the literature concerning the ability of lower-limb-worn IMUs to assess external load and additional kinematic/kinetic parameters amongst competitive athletes. To address this gap, the present systematic review aims to examine the extent to which IMUs have been employed in competitive sporting environments to monitor lower limb external load. Additionally, a secondary goal is to inform practice and future research in these areas.

## 2. Methods

### 2.1 Search strategy

This review was conducted following the guidelines proposed by Rico-González, Pino-Ortega (20). These guidelines take an adapted approach to the widely recognized Preferred Reporting Items for Systematic Reviews and Meta-analysis (PRISMA) protocol (21) to optimize the systematic review reporting in the field of sport science. Our review protocol was registered on 3/25/2024 on PROSPERO (ID: CRD42024526056).

Sources were initially searched with the earliest record through March 2024. A comprehensive search was conducted across multiple electronic databases, including PubMed, Web of Science, MEDLINE, Scopus, and SPORTDiscus, which have been identified as the most frequently used sources in sport science literature (20). No language restrictions were applied during the initial search; however, studies that were not available in full-text English were subsequently excluded.

The final search strategy utilized the following Boolean logic:

((Sport* OR Athlete* OR Team OR Player* OR Practice OR Training OR Game* or Match*) AND (Wearable OR Inertial Measurement Unit OR Microtechnology OR Sensor* OR IMU OR Microsensor OR Magnetometer OR Gyroscope OR Acceleromet*) AND (Lower extremity* OR Lower Limb* OR Shank OR Tibial OR Ankle OR Foot OR Lower Body) AND (Runn* OR work* OR Training Load OR Monitoring OR Load OR Step*))

### 2.2 Screening and Eligibility Criteria

Two independent reviewers (AL and JS) screened the search results against the predetermined eligibility criteria. Reviewers were not blinded to the titles or authors of the publications. Any disagreements were resolved through discussion and consensus, without requiring the involvement of a third researcher. Full texts of relevant studies were retrieved and independently reviewed using the same procedures. Additionally, reference lists of included studies were screened to identify any missing relevant articles. The identification, screening, and selection process is depicted in Figure 1.

**Figure 1.**
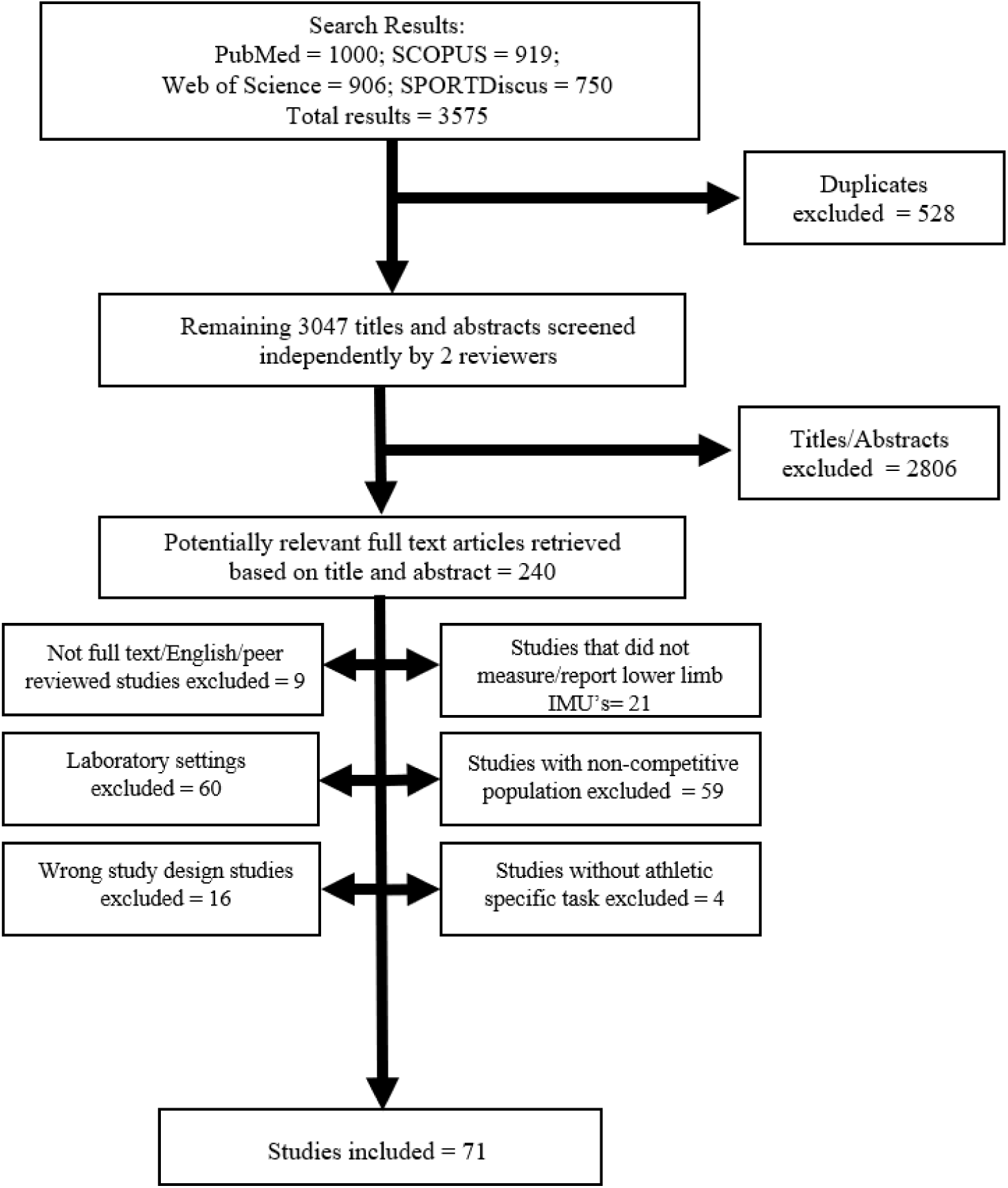
PRIMSA study selection process flow chart.

Studies were eligible for inclusion if they examined the use of IMUs worn or mounted below the knee in a natural sporting environment (e.g., field, pitch, court, track) and used a sample of competitive athletes participating in tasks related to their sport. “Competitive athlete” was defined according to the Bethesda Conference as ‘one who participates in an organized team or individual sport that requires regular competition against others as a central component, places a high premium on excellence and achievement, and requires some form of systematic and usually intense training’ (22). Studies were excluded if they (1) did not clearly define competitive athlete (e.g., ‘trained adults’, ‘recreational runners’); (2) evaluated non-human data; (3) were not available in English full text; (4) reported only laboratory-based monitoring or unauthentic (i.e. did not occur during normal team training or competition) sport drills. A risk of bias assessment was not conducted, as this review is descriptive and does not report or discuss effects, associations, or prevalence measures.

## 3. Results

### 3.1 Publication information

Since the year 2010 it is evident there is a clear rise in the usage of IMUs in sport (Fig). Of the 71 studies included in this review, all but one (23) were published after 2009. More recently there has been a surge in interest as 50 papers (70%) were published in the year 2020 or later.

A wide variety of journals were found to carry publications related to IMUs in sport with a total of 40 unique journals included. Of these, the most common included Sensors (12.7%; 9 studies); Journal of Sport Sciences and Sports Biomechanics (5.6%; 4 each); European Journal of Sport Science, IEE, Journal of Biomechanics, Science and Medicine in Football, and Medicine & Science in Sports & Exercise (4.2%; 3 each).

The country of publication was determined by the lead author’s contact information or the designated Institutional Review Board if no author information was provided. Information from a total of 18 distinct counties were included in this review. The majority of research was published out of European nations (70%, 50 studies) with a lesser contribution from North America (11 studies; 15%), Australia (8%, 6 studies), and Asia (6%, 5 studies). More than half of the research included in this review came from four countries, including the United Kingdom (14%), Spain (14%), the United States (13%), and Switzerland (11%).

Of the 71 studies included in this review, there were consistent themes across all aims of studies. The most common themes were the assessment of stride and/or foot spatiotemporal parameters in sport as this was the primary intention of 26 studies extracted. Spatial parameters refer to changes in orientation, including stride length, stride height, angular displacement, etc. Temporal parameters typically include time intervals, phase detection, and/or frequency of movements (24). A few common examples of this included detecting ground contact time, flight time, and stride cadence. Secondly, a similar quantity of studies (25) investigated validity and/or reliability. A significant portion of these papers aimed at validating the detection of unique events, also referred to as human-activity recognition (HAR), in the context of precise sporting actions. These include quantification of swimming kick-load (25, 26), distinct movements during ballet training (27), fast bowling phases (28), ball kicks/punts (29–31), and tennis serve types (32). Lastly approximately 20 articles aimed to quantity external load. In the context of this review, external load includes all quantification of biomechanical stressors of the lower limb, commonly including the magnitude and/or sum of accelerations and/or steps.

### 3.2 Participant demographics

The number of participants ranged broadly from a single-subject case study on an elite trail runner’s stride parameters (33) to multi-club investigations including 293 professional females soccer players (34) comparing external load across competitive levels. While the median number of participants was 15, only 10 of 71 (14%) publications explicitly justified their sample via power analysis (35–44). Due to the nature of this review targeting competitive athletics, it is inferred that most authors leveraged a convenience sample, while only two studies explicitly specified this (31, 45).

Over half of the included publications focused on male athletes (52%; 37), while approximately a third examined both males and females (32%; 23). Only a small proportion studied exclusively female athletes (6%; 4), and some did not specify the participants’ sex (10%; 7).

The primary categorization of sport is delineated by Stefani (47) where the competitive nature is defined as independent, or team object-based. The present review examined 38 independent, 32 team object sports, and 1 combination that included athletes from both individual (track and field) and team object (soccer) sports (48).

The next level of taxonomy was adopted from McKay, Stellingwerff (49) who segregated the sports based on the physiological, tactical, and skill demands. These included endurance (16), middle distance (6), speed/strength (10), combat (2), and ‘other’ (4) which included dance and gymnastics. The performance caliber tiers established by McKay, Stellingwerff (49) utilize a 0-5 tiered system. In this, 0 is sedentary, 1 is recreational, 2 is trained/developmental, 3 is highly trained/national, 4 is elite/international, and 5 is world-class/Olympic. The breakdown of these is shown in Figure 3. Two-thirds of the studies observing team sport athletes examined 3^rd^ and 4^th^ tier athletes, 25% and 41%, respectively. The most studied team object sports were soccer (15), basketball (5), and Australian Rules Football (AFL)/rugby (4). The most independent sports studies included distance running (12; >5000 m) and sprinting (5; ≤ 400m).

**Figure 2.**
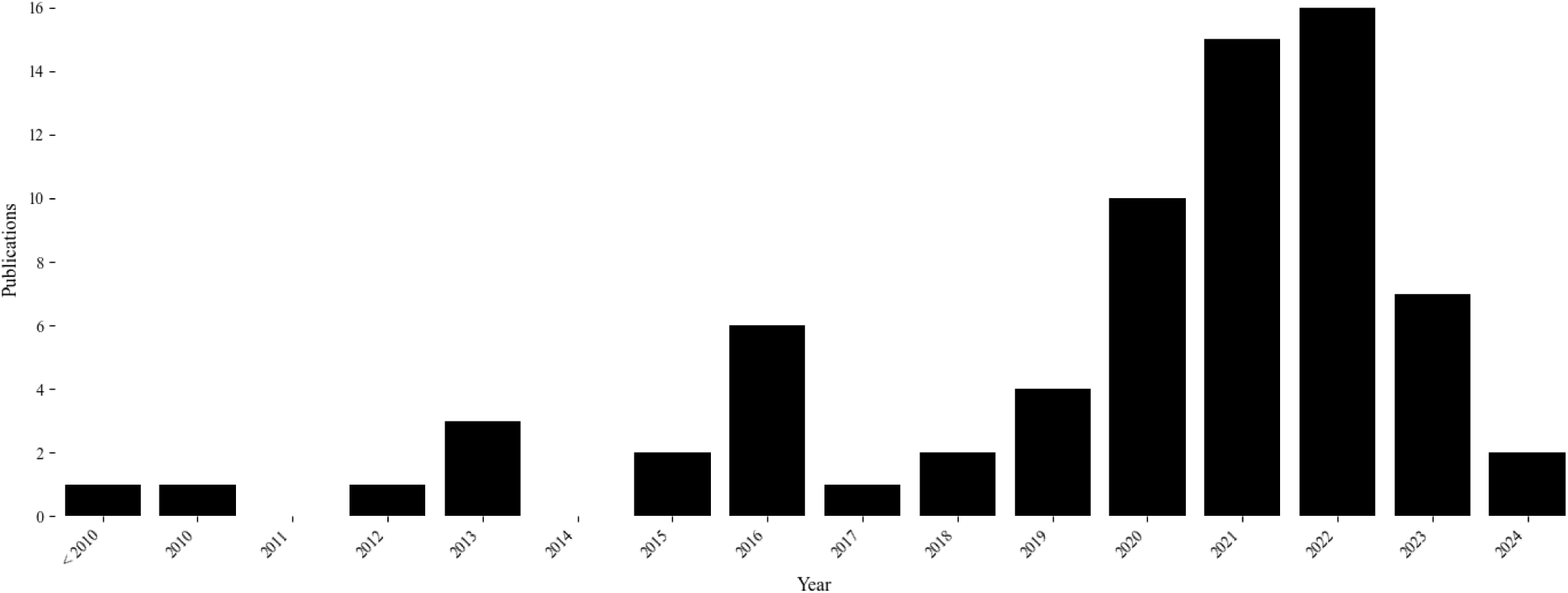
Research evolution of publications by year.

**Figure 3.**
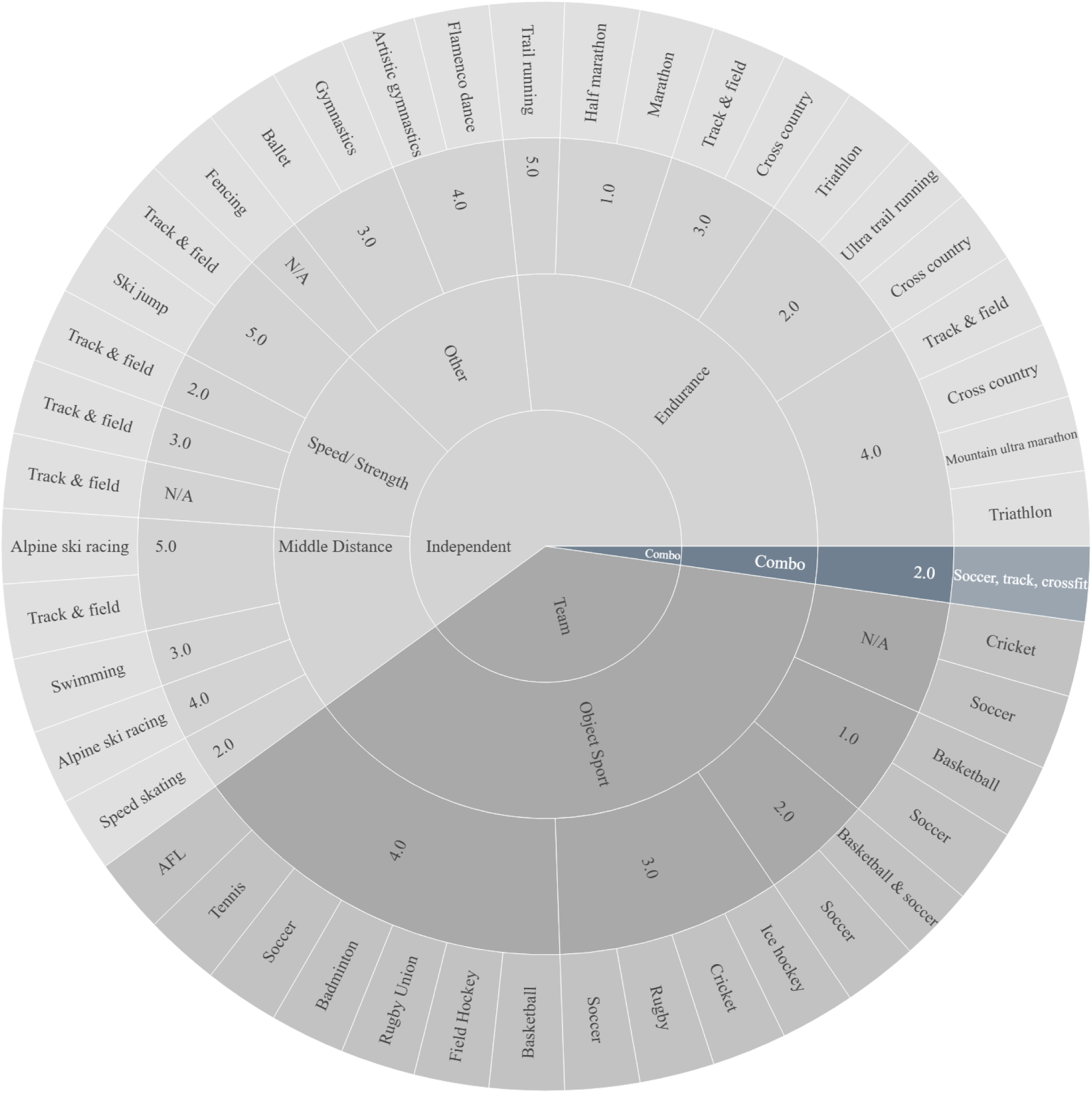
Sunburst plot of independent/team participation, sporting demands, performance caliber, and sport.

The independent athletes, however, had a more even distribution across all calibers, with the largest portion of research examining 3^rd^ tier athletes. The single combination/mixed sample publication studied a 2^nd^ tier participant group. The largest relative portion of individuals were endurance athletes (42%; 16) including a variety of distance-running sports related to marathon (35, 50–54), cross country (15, 36, 45, 55), and triathlon (25, 56).

### 3.3 Device specifications

Overall, a total of 25 device manufacturers and 35 sensor models were reported (Table 1). The level of detail regarding device specifications varied widely across all publications. In the event certain data was not provided by the authors, an internet search was completed to augment missing device specifications when possible. While five research-grade sensors did not specify the device sampling rate, 16 of the remaining 30 (53%) devices were operated between 250-1600 Hz. Median dynamic ranges of the internal components include ± 16g accelerometer, ± 2,000°/s gyroscope, and 1,200μT magnetometer. The median IMU size is 1225 cm^3^, which coincides with a device with the subtle dimensions of 3.5×3.5×1 cm. Across all devices, the median battery life reported was eight hours. The commercially manufactured IMeasureU Blue Thunder and Blue Trident devices were the most commonly deployed, comprising 14% (10 studies) of all publications in this review.

**Table 1.** Devices and specifications.

| Ref | Manufacturer | Model | L x W x H (mm) | Mass (g) | 3-axis Sampling Rate (Hz) | Accelerometer Dynamic Range (+/- g) | Gyroscope Dynamic Range (°/sec) | Magnetometer Dynamic Range (μT) | Battery |
| --- | --- | --- | --- | --- | --- | --- | --- | --- | --- |
| (30, 38, 45, 50, 53, 57, 58) | IMeasureU | Blue Thunder | 40x30x15 | 12 | 1000 | 16 | 2000 | 1200 | 6 h |
| (6, 7, 14, 59-61) | RealTrack Systems | WIMU Pro |  |  | 1000 | 16 |  | 1000 | 10 h |
| (48, 62, 63) | Invensense | MPU-9150 | 4x4x1 |  | 500 | 16 | 2000 | 1200 |  |
| (29, 31, 34, 64, 65) | PlayerMaker | N/A | 32x21x5 | 6 | 1000 | 16 | 2000 | 1200 |  |
| (51, 66, 67) | Xsense Technologies | MTw Awinda | 34.5x57.8x14.5 | 27 | 1000 |  | 1200 | 120 | 6 h |
| (39, 40, 42) | Biometrics ltd | ACL 300 | 19x12.7x10.9 |  |  | 10 |  | 1000 |  |
| (68-71) | GaitUp | Physilog 4 | 50x37x9.2 | 19 | 500 | 16 | 2000 | 1000 | 8 h |
| (35, 52, 72) | GaitUp | Physilog 5 | 47.5x26.5x10 | 11 | 512 | 16 | 2000 |  |  |
| (41, 73, 74) | IMeasureU | Blue Trident | 42x27x11 | 9.5 | 1600 | 16 | 2000 | 4900 | 12h |
| (75, 76) | Catapult Innovations | MinimaxX |  |  | 100 |  |  |  | 8 h |
| (77, 78) | Delsys | Trigno | 27x37x13 | 14 | 963 | 16 | 741 | 222 | 14 h |
| (33, 79) | HiKoB | Agile Fox |  |  |  |  |  |  |  |
| (36, 55) | Polar | S3+ |  |  | 1 |  |  |  | 8 h |
| (15, 80) | RunScribe | Plus | 32x25x7.5 | 15 | 500 | 16 | 2000 | 1200 | 2 h |
| (27) | ActiGraph | GT9x Link | 35x35x10 | 14 | 1000 | 16 | 2000 | 4800 |  |
| (81) | ActiGraph | Link | 35x35x10 | 14 | 100 | 16 | 2000 | 4800 | 9 d |
| (82) | Analog Devices | ADXL 78 |  | 35 | 2400 |  |  |  |  |
| (56) | APDM | Opal |  |  | 128 |  | 2000 |  |  |
| (28) | Bosch | Smart sensor | 5.2x3.8x1.13 |  |  | 14 | 2000 |  |  |
| (26) | Cometa | N/A |  |  | 285 | 16 | 2000 |  |  |
| (44) | Invensense | MPU-9255 | 44x30x8 |  | 100 | 16 | 2000 |  |  |
| (37) | Delsys | Trigno |  |  |  | 9 |  |  | 6 h |
| (43) | dorsaVi Ltd | N/A |  |  | 200 |  |  |  | 12 h |
| (83) | ErgoTest Innovation | N/A |  | 20 | 200 | 16 | 2000 | 2500 |  |
| (32) | Excel | EXLs3 | 54x33x14 | 22 | 200 | 16 | 2000 |  |  |
| (25) | Garmin | Forerunner 735 XT |  |  | 25 | 1 |  |  | 12 d |
| (84) | HuCE-microLab | PARTwear | 38x37x8 |  | 1000 | 16 |  |  |  |
| (23) | IC Sensors | Model 3031 |  | 2.5 | 1200 | 20 |  |  |  |
| (85) | Noitom Technology | AxisNeurono | 28x16x12 | 4.1 | 120 |  |  |  | 200 h |
| (86) | Noraxon | MyoMotion | 37.6x52x18.1 | 34 | 200 | 16 | 400 |  | 3 h |
| (87) | Phillips Healthcare | Actical | 29x37x11 | 16 | 32 | 2 |  |  |  |
| (88) | Physilog | BioAGM |  | 100 | 500 | 10 | 1200 |  | 3 h |
| (89) | SABEL Labs | SABEL Sense | 55x30x13 | 23 |  |  |  |  |  |
| (90) | TalentPlayers | IMU |  |  | 100 |  |  |  |  |

**Table 2.**
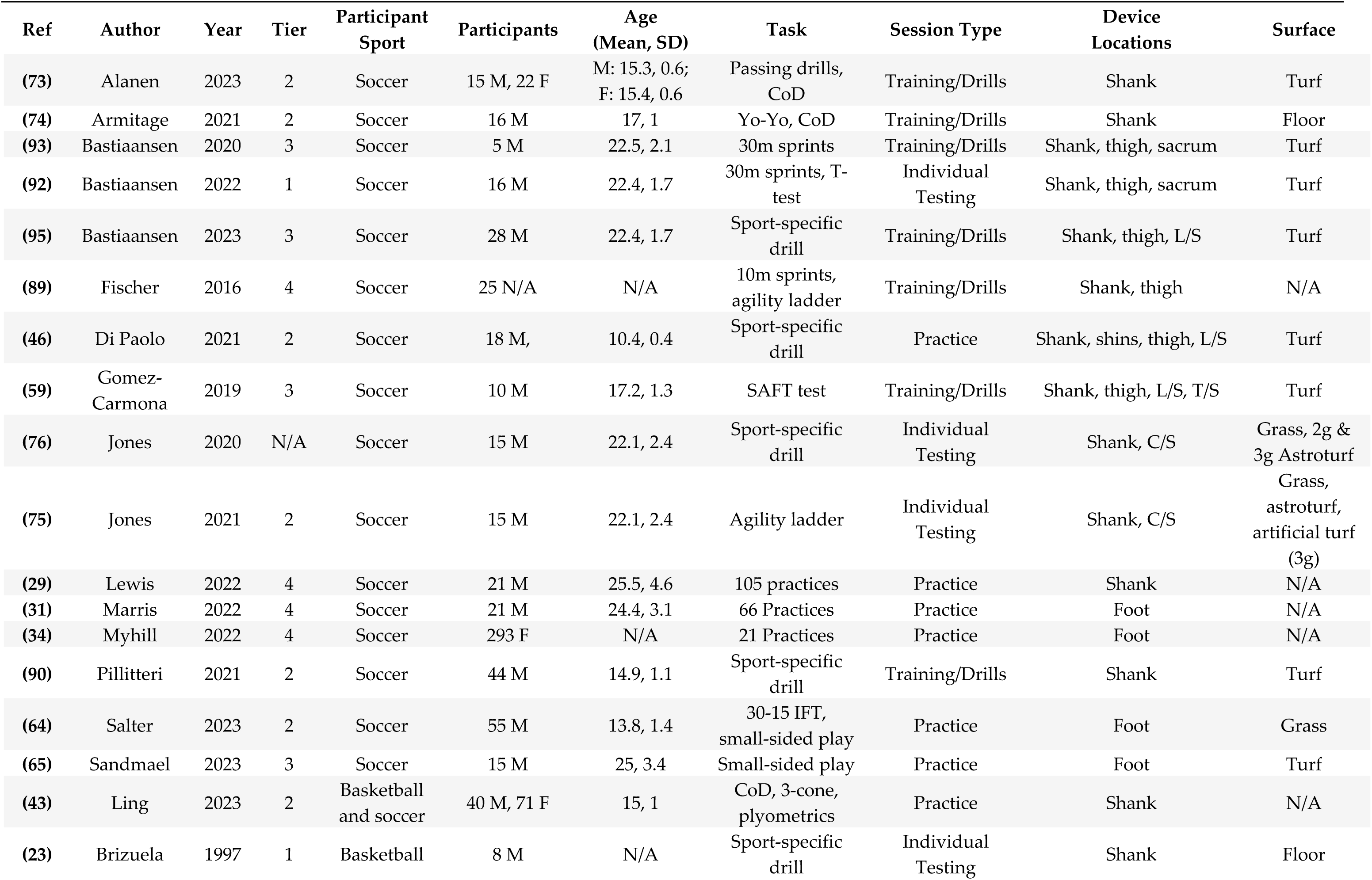
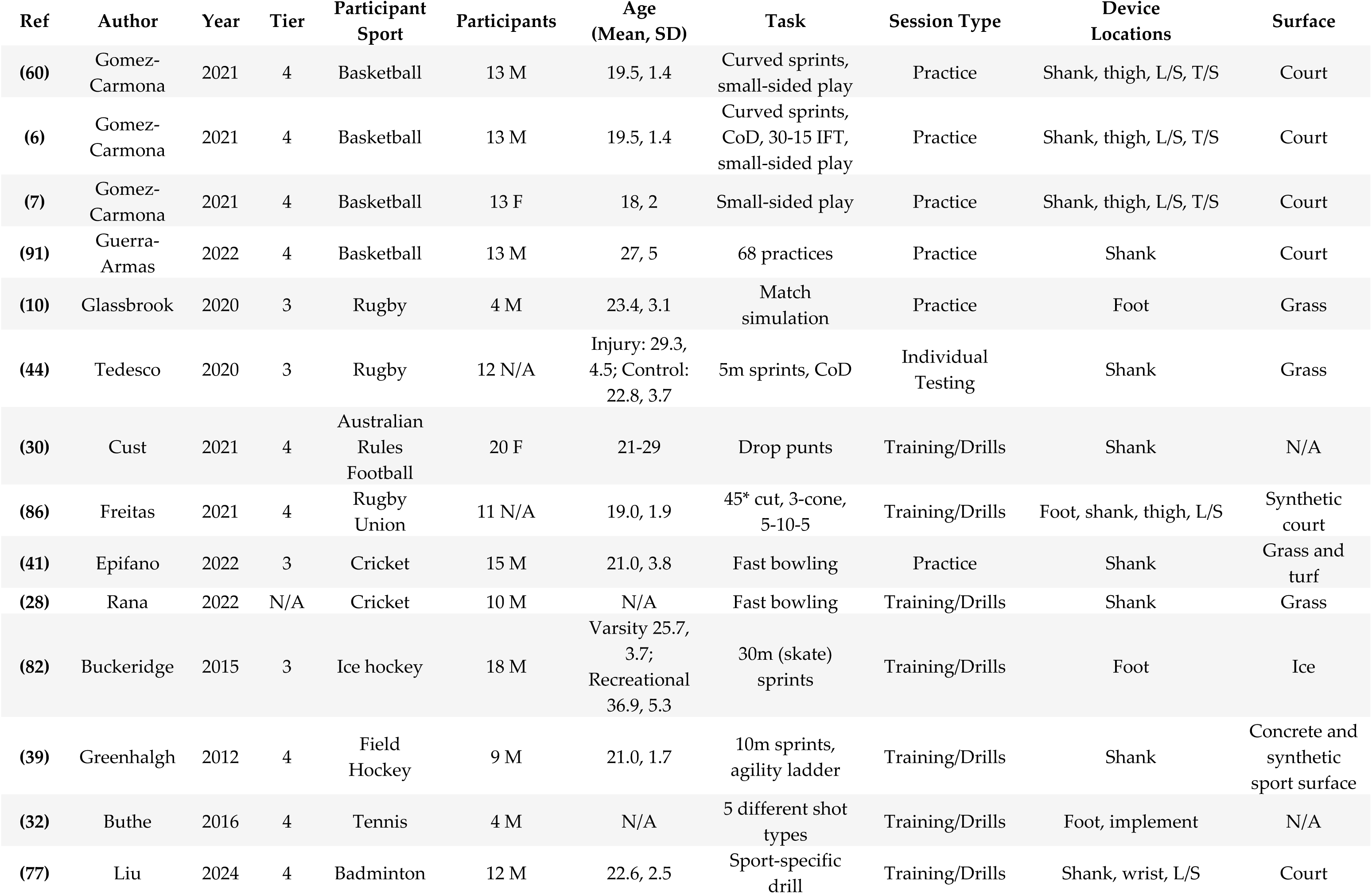
Team object sport athlete studies.

**Table 3.**
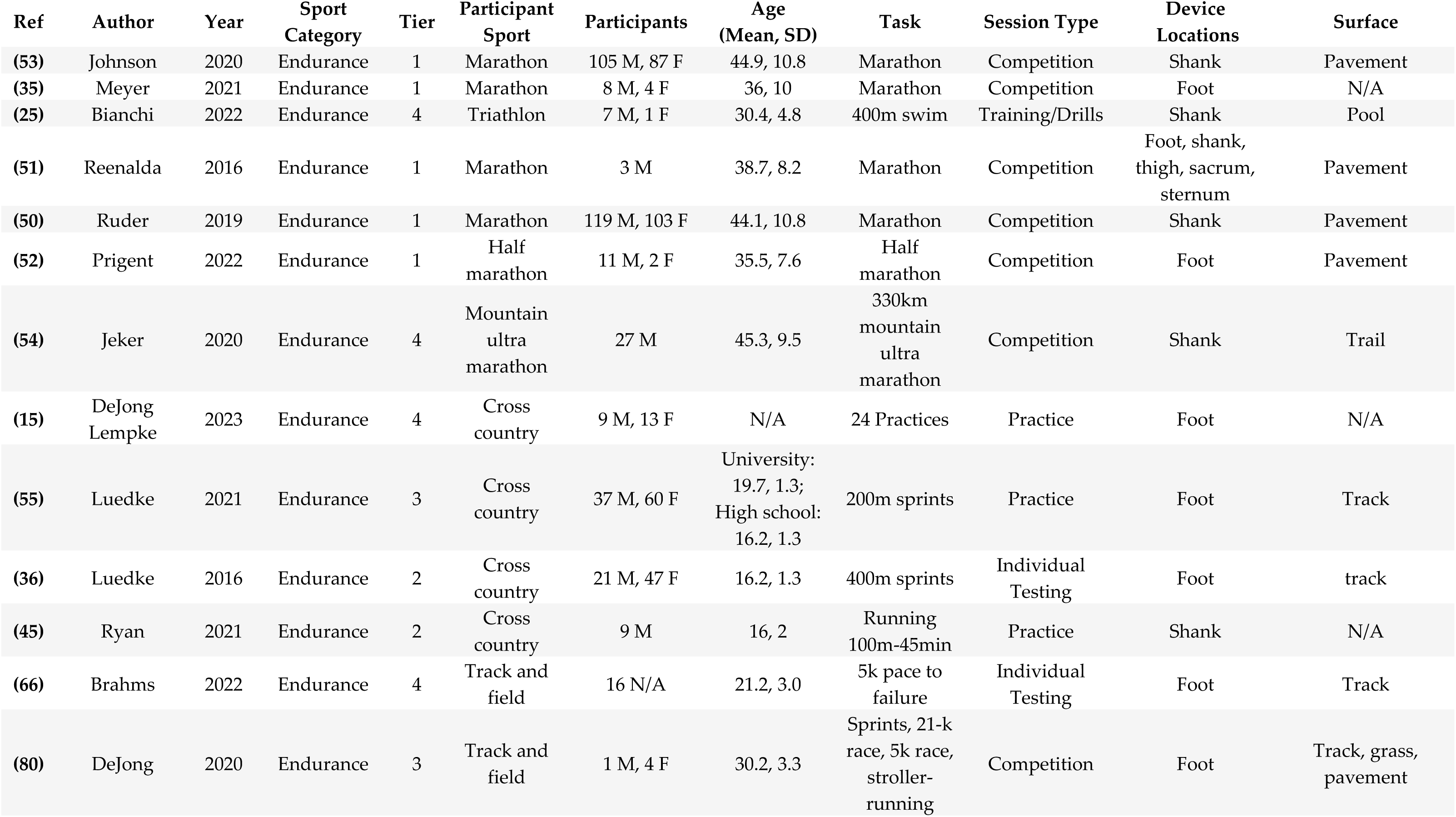
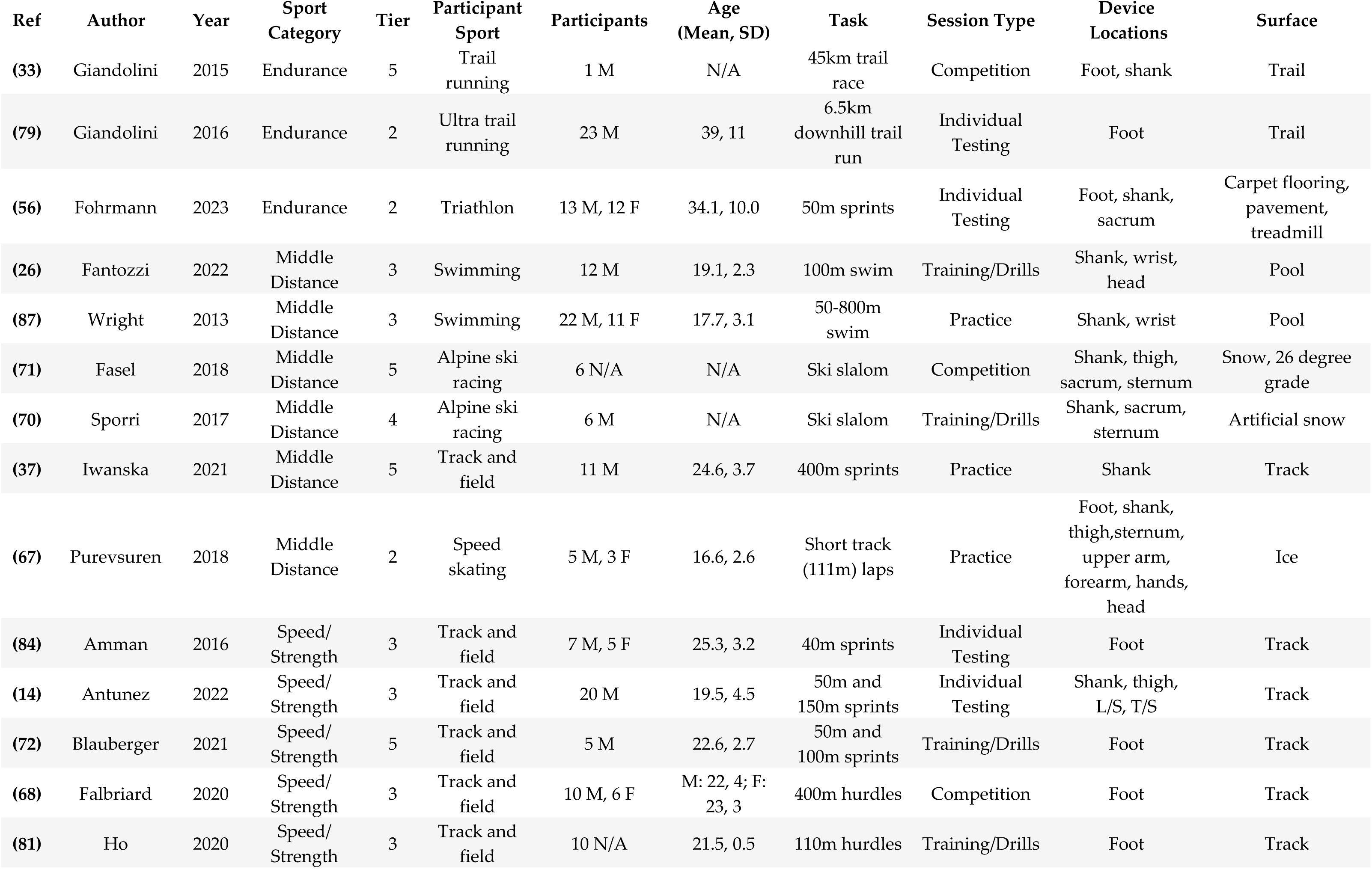
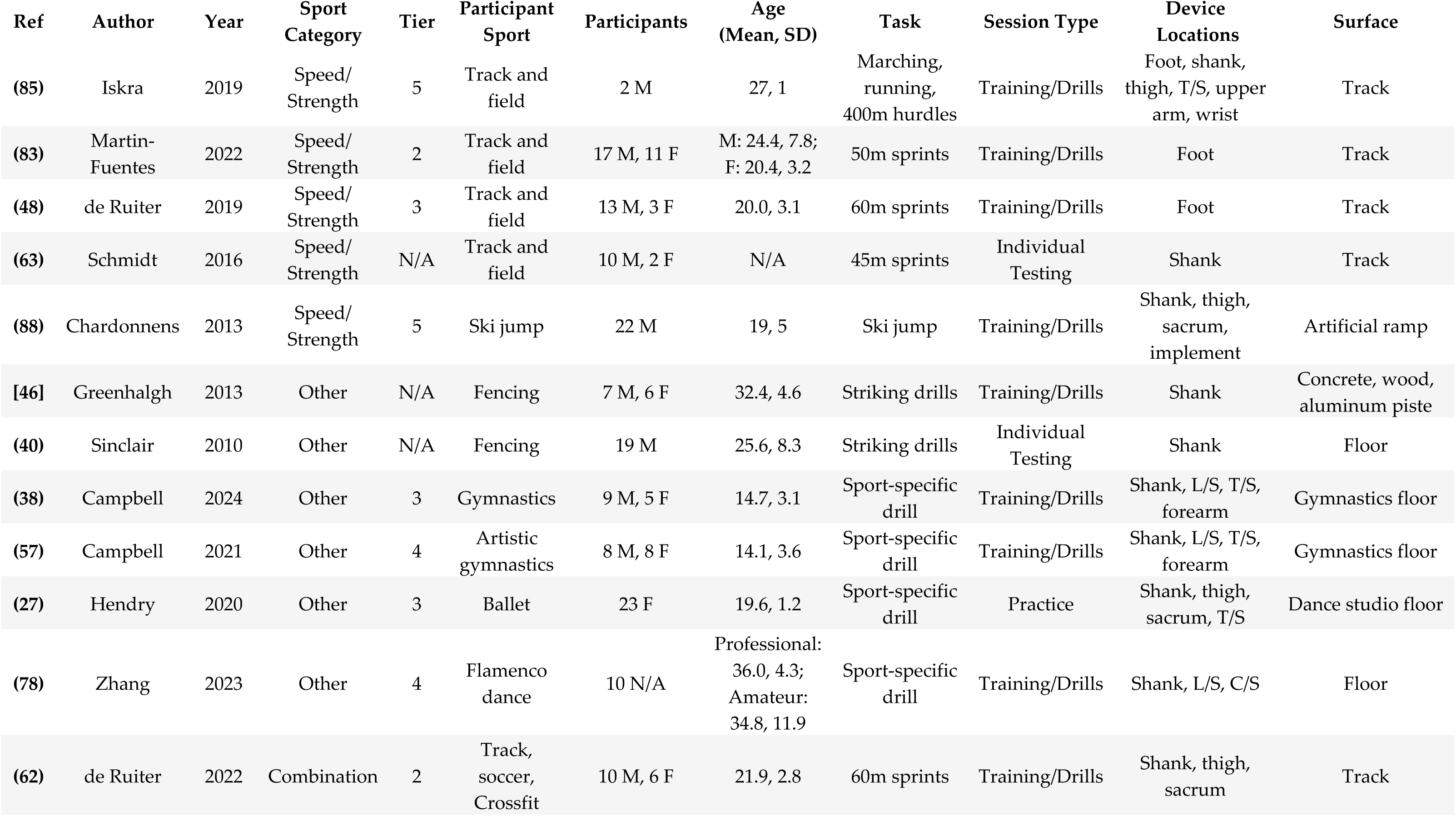
Independent sport athlete studies.

| Ref | Author | Year | Sport Category | Tier | Participant Sport | Participants | Age (Mean, SD) | Task | Session Type | Device Locations | Surface |
| --- | --- | --- | --- | --- | --- | --- | --- | --- | --- | --- | --- |
| (53) | Johnson | 2020 | Endurance | 1 | Marathon | 105 M, 87 F | 44.9, 10.8 | Marathon | Competition | Shank | Pavement |
| (35) | Meyer | 2021 | Endurance | 1 | Marathon | 8 M, 4 F | 36, 10 | Marathon | Competition | Foot | N/A |
| (25) | Bianchi | 2022 | Endurance | 4 | Triathlon | 7 M, 1 F | 30.4, 4.8 | 400m swim | Training/Drills | Shank | Pool |
| (51) | Reenalda | 2016 | Endurance | 1 | Marathon | 3 M | 38.7, 8.2 | Marathon | Competition | Foot, shank, thigh, sacrum, sternum | Pavement |
| (50) | Ruder | 2019 | Endurance | 1 | Marathon | 119 M, 103 F | 44.1, 10.8 | Marathon | Competition | Shank | Pavement |
| (52) | Prigent | 2022 | Endurance | 1 | Half marathon | 11 M, 2 F | 35.5, 7.6 | Half marathon | Competition | Foot | Pavement |
| (54) | Jeker | 2020 | Endurance | 4 | Mountain ultra marathon | 27 M | 45.3, 9.5 | 330km mountain ultra marathon | Competition | Shank | Trail |
| (15) | DeJong Lempke | 2023 | Endurance | 4 | Cross country | 9 M, 13 F | N/A | 24 Practices | Practice | Foot | N/A |
| (55) | Luedke | 2021 | Endurance | 3 | Cross country | 37 M, 60 F | University: 19.7, 1.3; High school: 16.2, 1.3 | 200m sprints | Practice | Foot | Track |
| (36) | Luedke | 2016 | Endurance | 2 | Cross country | 21 M, 47 F | 16.2, 1.3 | 400m sprints | Individual Testing | Foot | track |
| (45) | Ryan | 2021 | Endurance | 2 | Cross country | 9 M | 16, 2 | Running 100m-45min | Practice | Shank | N/A |
| (66) | Brahms | 2022 | Endurance | 4 | Track and field | 16 N/A | 21.2, 3.0 | 5k pace to failure | Individual Testing | Foot | Track |
| (80) | DeJong | 2020 | Endurance | 3 | Track and field | 1 M, 4 F | 30.2, 3.3 | Sprints, 21-k race, 5k race, stroller-running | Competition | Foot | Track, grass, pavement |
| (33) | Giandolini | 2015 | Endurance | 5 | Trail running | 1 M | N/A | 45km trail race | Competition | Foot, shank | Trail |
| (79) | Giandolini | 2016 | Endurance | 2 | Ultra trail running | 23 M | 39, 11 | 6.5km downhill trail run | Individual Testing | Foot | Trail |
| (56) | Fohrmann | 2023 | Endurance | 2 | Triathlon | 13 M, 12 F | 34.1, 10.0 | 50m sprints | Individual Testing | Foot, shank, sacrum | Carpet flooring, pavement, treadmill |
| (26) | Fantozzi | 2022 | Middle Distance | 3 | Swimming | 12 M | 19.1, 2.3 | 100m swim | Training/Drills | Shank, wrist, head | Pool |
| (87) | Wright | 2013 | Middle Distance | 3 | Swimming | 22 M, 11 F | 17.7, 3.1 | 50-800m swim | Practice | Shank, wrist | Pool |
| (71) | Fasel | 2018 | Middle Distance | 5 | Alpine ski racing | 6 N/A | N/A | Ski slalom | Competition | Shank, thigh, sacrum, sternum | Snow, 26 degree grade |
| (70) | Sporri | 2017 | Middle Distance | 4 | Alpine ski racing | 6 M | N/A | Ski slalom | Training/Drills | Shank, sacrum, sternum | Artificial snow |
| (37) | Iwanska | 2021 | Middle Distance | 5 | Track and field | 11 M | 24.6, 3.7 | 400m sprints | Practice | Shank | Track |
| (67) | Purevsuren | 2018 | Middle Distance | 2 | Speed skating | 5 M, 3 F | 16.6, 2.6 | Short track (111m) laps | Practice | Foot, shank, thigh, sternum, upper arm, forearm, hands, head | Ice |
| (84) | Amman | 2016 | Speed/Strength | 3 | Track and field | 7 M, 5 F | 25.3, 3.2 | 40m sprints | Individual Testing | Foot | Track |
| (14) | Antunez | 2022 | Speed/Strength | 3 | Track and field | 20 M | 19.5, 4.5 | 50m and 150m sprints | Individual Testing | Shank, thigh, L/S, T/S | Track |
| (72) | Blauberger | 2021 | Speed/Strength | 5 | Track and field | 5 M | 22.6, 2.7 | 50m and 100m sprints | Training/Drills | Foot | Track |
| (68) | Falbriard | 2020 | Speed/Strength | 3 | Track and field | 10 M, 6 F | M: 22, 4; F: 23, 3 | 400m hurdles | Competition | Foot | Track |
| (81) | Ho | 2020 | Speed/Strength | 3 | Track and field | 10 N/A | 21.5, 0.5 | 110m hurdles | Training/Drills | Foot | Track |
| (85) | Iskra | 2019 | Speed/Strength | 5 | Track and field | 2 M | 27, 1 | Marching, running, 400m hurdles | Training/Drills | Foot, shank, thigh, T/S, upper arm, wrist | Track |
| (83) | Martin-Fuentes | 2022 | Speed/Strength | 2 | Track and field | 17 M, 11 F | M: 24.4, 7.8; F: 20.4, 3.2 | 50m sprints | Training/Drills | Foot | Track |
| (48) | de Ruiter | 2019 | Speed/Strength | 3 | Track and field | 13 M, 3 F | 20.0, 3.1 | 60m sprints | Training/Drills | Foot | Track |
| (63) | Schmidt | 2016 | Speed/Strength | N/A | Track and field | 10 M, 2 F | N/A | 45m sprints | Individual Testing | Shank | Track |
| (88) | Chardonnnens | 2013 | Speed/Strength | 5 | Ski jump | 22 M | 19, 5 | Ski jump | Training/Drills | Shank, thigh, sacrum, implement | Artificial ramp |
| [46] | Greenhalgh | 2013 | Other | N/A | Fencing | 7 M, 6 F | 32.4, 4.6 | Striking drills | Training/Drills | Shank | Concrete, wood, aluminum piste |
| (40) | Sinclair | 2010 | Other | N/A | Fencing | 19 M | 25.6, 8.3 | Striking drills | Individual Testing | Shank | Floor |
| (38) | Campbell | 2024 | Other | 3 | Gymnastics | 9 M, 5 F | 14.7, 3.1 | Sport-specific drill | Training/Drills | Shank, L/S, T/S, forearm | Gymnastics floor |
| (57) | Campbell | 2021 | Other | 4 | Artistic gymnastics | 8 M, 8 F | 14.1, 3.6 | Sport-specific drill | Training/Drills | Shank, L/S, T/S, forearm | Gymnastics floor |
| (27) | Hendry | 2020 | Other | 3 | Ballet | 23 F | 19.6, 1.2 | Sport-specific drill | Practice | Shank, thigh, sacrum, T/S | Dance studio floor |
| (78) | Zhang | 2023 | Other | 4 | Flamenco dance | 10 N/A | Professional: 36.0, 4.3; Amateur: 34.8, 11.9 | Sport-specific drill | Training/Drills | Shank, L/S, C/S | Floor |
| (62) | de Ruiter | 2022 | Combination | 2 | Track, soccer, Crossfit | 10 M, 6 F | 21.9, 2.8 | 60m sprints | Training/Drills | Shank, thigh, sacrum | Track |

All studies included in this review studied IMU(s) below the knee. Of these, most had devices worn/mounted on the shank (44), with fewer publications having sensors at the foot (21) or shank and foot (6). Almost half the studies (30) did observe sensors at multiple locations superior to the lower limb. The most common sites included sacrum/lumbar (21), thigh (18), cervical/thoracic (12), upper arm/forearm (5), wrist/hand (5), sternum (4), head (2), and implements (2). At least 79% (56) of all studies used devices bilaterally.

### 3.4 Sporting task

The majority of study designs followed a cross-sectional methodology (77%) assessing athletes at one point in time, while few deployed a longitudinal monitoring approach using a cohort design (15%). The remaining 8% was comprised of three case studies and two case-control designs. Of these team-object sport longitudinal studies, 75% (8 of 12) monitored full practice sessions or small-sided games mimicking match-like conditions.

Unless explicitly specified by the authors, it was inferred whether the data was collected during competition, practice, training/drills, or individual testing sessions. The team-object sport publications predominantly monitored athletes most during specific training/drills (43%; 14) and practice (40%; 13), with fewer individual testing sessions (16%, 5) dedicated solely to data collection. The most common tasks among team-object sport publications were sport-specific drills (38%, 12 of 32 papers). Of these were soccer-specific change of direction and passing drills (46, 73, 76, 90), fast bowling (28, 41), basketball-specific drills (23), badminton shots (77), tennis shots (32), and Australian Rules Football kicks (30). Following tasks specific to their sport, athletes were monitored during more general change of direction and/or agility drills (6, 39, 43, 44, 60, 73–76, 86, 89), small-sided games/practices (6, 7, 29, 31, 34, 58, 60, 64, 65, 91), and short sprints ranging from 5-30m (39, 44, 82, 89, 92, 93).

Individual sport athletes were most commonly monitored during training/drills (34%; 13) followed by competition (26%; 10), individual testing (21%; 8), and practice (18%; 7). Most all individual sport observations were running-based with some exceptions including swimming (25, 26, 87), ski (70, 71, 88), fencing strikes (40, 94), dance (27, 78), gymnastics (38, 57) and speed skating (67). The most common independent running athletes were endurance-oriented with IMU assessments ranging from 100m (45) to 330km (80), with marathon-like distances most common (33, 35, 50–53). Independent middle distance runners performed 400m sprints (37) and independent speed/strength running athletes tasks included sprints/hurdles between at distances from 40-400m (14, 48, 63, 68, 72, 81, 83–85).

If applicable, it was extracted whether the studies were performed indoors or outdoors since IMUs allow more flexibility in not relying on GNSS receivers or LPS beacons. While 13% (9) papers failed to report on location, approximately half (52%; 32) of all publications monitored athletes outdoors. The remainder (35%, 24) collected data indoors or a combination of indoors and outdoors (1%; 1) (56).

The surface for athletic tasks was recorded if the authors explicitly stated such information or it could be inferred based on additional information throughout the publication (photos, indoor/outdoor, etc.). A total of 14% (10) were not reliably identified and left as non-applicable and an additional 11% (8) noted tasks on multiple different surfaces. Among the most common surfaces included synthetic track (15), artificial turf (12), grass (9), and concrete/pavement (8).

## 4. Discussion

### 4.1 General trends

The primary purpose of this review was to synthesize the use of lower limb monitoring with IMUs in competitive sport. This manuscript provides an overview of publication information, participant demographics, device specifications, and sporting tasks that are examined in the literature to date. Several reviews have shown that wearable usage in sport has rapidly increased over recent years (17, 24, 96–98). However, this investigation offers novel insights into the sporting contexts in which IMUs are utilized to capture external load data specific to the lower limbs. As practitioners implement more targeted monitoring strategies, new opportunities arise to refine training prescriptions and better distinguish whole-body external load from the specific demands on the lower limbs, reducing the risk of overlooked external stress.

While it was previously established that lower limb-worn IMUs are most often utilized indoors on treadmills, this review garnished additional implementation in sport beyond laboratory settings as previous reviews have called for (17, 99, 100). At this time is recognized that there is a marked increase in the usage of lower limb assessments with IMUs in competitive environments. The present review shows that most of these assessments are typically carried out during training/practice rather than competitive matches/games. The lack of IMU deployment in team-object sport matches could be explained by certain professional leagues prohibiting the use of such monitoring approaches during gameplay as there is a conflict of interest with league-sponsored technologies established in collective bargaining agreements (101, 102). This is likely why individual endurance athletes used these technologies during competitive races at a higher rate since no such agreement prohibits such usage (33, 35, 50–54).

The publications outlining the implementation of lower limb IMUs are notably different when comparing team and independent athletes. There was a higher proportion of publications from team sport athletes in the 4th tier (and Division I university athletes), while publications involving individual athletes reported more participants from lower level university athletics, developmental, and recreational populations. There are several potential reasons for this, including logistical constraints that play a role in these differences, including the challenges in feasibility monitoring larger groups or the financial burden associated with some technology use. Higher level athletes may also consider this data as proprietary or a competitive advantage, creating an unwillingness to share it publicly through publications. This could explain why this review found no team sport studies published with tier 5 (Olympic or world-class) athletes. The assumed recruitment strategies of these groups differ quite significantly as well. Competitive endurance runners can be contacted to participate in research by providing their information during local races, such as marathon registration (50, 53). Team sport research, however, is typically limited to smaller convenience sampling. Mitchell, Ratcliff (103) recently wrote that the recruitment of 3^rd^ tier and higher (team sport) athletes is most often done via an internal gatekeeper that is affiliated with the team/club, where this was most often a contributing author. A previously suggested means to facilitate more research in these environments is embedding sports science researchers who can identify and work within the circumscriptions of such settings (104, 105).

Differentiating the caliber of athletes and varying competitive levels has been recommended as best practice in sport science research (49). While this review did not include a metanalytic approach, there were evident themes regarding differences in performance caliber worth considering. In endurance sports, higher running velocities are associated with greater tibial acceleration, reflecting the increased external load demands on advanced athletes (55, 66, 106). Peak tibial acceleration, often termed tibial shock (50), provides insight into lower limb demands during training and competition. While the strength of this association varies in the literature, vertical tibial acceleration is at least moderately correlated with ground reaction forces (GRFs) during running providing kinetic inferences (107–110). However, external load— represented by the magnitude and volume of accumulated GRFs—should not be directly equated with internal forces experienced by the lower limb (18, 111). Although recent criticism questions the direct use of tibial acceleration to infer kinetic data regarding tibial load (112), a consistent positive relationship between tibial acceleration and running speed remains (106). Likewise, additional kinematics differ in elite populations, including elevated cadences (55) and longer stride lengths at similar cadences (18, 66). In team sports there are several noted biomechanical differences across calibers outside of merely tibial acceleration including greater ranges of motion and angular velocities during skating drills (82) and soccer (95). These data highlight the inherent differences in external load across performance levels, emphasizing the need for tailored training prescriptions based on an athlete’s performance caliber.

There has been an effort to distinguish the onset of fatigue using IMUs during cyclical exercise (113). As fatigue sets in during running, shock attenuation decreases, and peak tibial acceleration increases. It is noted that across distance running events, peak impact acceleration tends to rise over time accompanied by a 14% reduction in stride length and a 9% increase in contact time (66). Additionally, stride height and stride frequency decline over the course of a race (54), with changes in spatiotemporal stride parameters, including an increased duty factor (35). Stride length alterations also occur but appear to be nonuniform, as seen in a marathon where stride length decreased in two runners but increased in one (51). Similarly, in elite soccer players, changes in mediolateral ankle acceleration has been identified as a marker of fatigue (89). These findings highlight the individualized nature of fatigue-related biomechanical changes, emphasizing the need for athlete-specific monitoring and intervention strategies.

### 4.2 Device considerations

While lower limb-worn IMUs have been identified to create several advantages throughout this review, there remains no clear device gold standard for athlete monitoring (100). A key consideration previously identified is the acknowledgment that load is site-specific to the anatomical location of the wearable deployed where distally worn IMUs will present elevated values (6, 7, 11, 13, 57, 59, 60, 75, 77, 92). Also, when these devices are worn at the distal shank there is significantly greater between-subject variability compared to the thoracic and lumbar areas (6, 56). Likewise, lower limb-worn IMUs display a greater relative percentage of mediolateral and anterior-posterior external load (75). This underscores the value of triaxial accelerometry in uncovering how external load is distributed across different planes in different sporting tasks. Additionally, the athlete’s internal load may be better inferred as the external load at the shank was more highly correlated with relative heart rate reserve than when worn at the trunk (114). The information gathered from the lower limb is additive to understanding how movement strategies differ among individuals, where whole-body external load may be less sensitive to such detection.

Differences between limbs can be detected when devices are worn bilaterally, although several studies opted to assess only one limb (23, 25, 28, 30, 33, 36, 40, 50, 55, 66, 79, 82, 94). The assessment of athletic asymmetries has been somewhat common practice while it is recognized that interlimb differences may vary at an individual and/or task-specific level (115–118). It is noted that curved running imposes asymmetrical kinematic demands with greater acceleration magnitudes garnished from the outside leg (60, 119–121) as compared to linear running. Additionally, the inside leg tends to have longer GCTs with smaller GRFs (119, 120). Running-based tasks in sport present very little exclusively linear movement, imposing inherent interlimb differences that may not be functionally advantageous. Some runners are shown to possess differing levels of efficiency in curved running, providing opportunities to bolster this attribute in training with the appropriate monitoring (14). For a given athletic task, practitioners may consider the upper limits of tolerable asymmetry related to the ecological norms of the sport (106). For example, performance on a 200m track is typically slower than 400m (distance controlled) due to smaller radii likely imposing greater levels of interlimb asymmetry (120). Also, runners with a history of lower extremity injuries have presented with longer GCTs on such limbs compared to their healthy counterparts (44, 122). Preemptively, it has been shown lower limb-worn IMUs may detect spatiotemporal differences in distance runners leading up to injury (80). While healthy human movement is rarely perfectly symmetrical (58, 85, 106), monitoring interlimb kinematic differences can provide the opportunity to mitigate unwarranted external loads and mitigate future injury (93).

Previous research has called for enhanced ease of use and easily interpretable metrics to elevate the utilization of IMUs in practice beyond research settings (17, 24). While this review did not report on specific metrics among all studies, there is a noted level of heterogeneity that is contingent upon the devices deployed and/or analysis/methodology performed by each investigator. For example, several papers did not report on metric outputs but rather analyzed all data in its raw form with custom scripts (50, 51, 53). Additionally, when researchers are utilizing custom-made devices there is potential for bespoke opportunities with custom programs/scripts (44) which can pose challenges to reproducing findings. This can be augmented with more open-source documentation similar to recent recommendations on how to identify initial and terminal contact of the stance phase based on IMU location (123). Caution should be expressed as this does not necessitate manufacturers to leverage these publications to advance consumer-grade sports products. HAR literature also shows promise to automate certain sporting tasks via supervised classification algorithms to include the identification of different tennis shot types (32), AFL kick types (30), and bowling phases (41). It is worth noting that some HAR events, beyond the spatiotemporal metrics of the gait cycle, still require multiple sensors and reduce feasibility in sport (124). While these advancements have shown promise, they continue to demonstrate the sustained divide between what can be accomplished in research and what is widely deployable in the natural sporting ecology.

### 4.3 Practical Recommendations

Monitoring external load provides value by characterizing drills, practices, and competition (125). As practitioners accumulate knowledge surrounding the normative demands for such sporting events, this information can be leveraged to reinforce feedback loops, ultimately optimizing the training prescriptions and athlete outcomes. Site-specific IMU placement can be done with commercially available devices to quantify loads that are more precise to bodily regions of interest, given the demands of differing sporting tasks. Practitioners refining their athlete monitoring programs should consider leveraging lower-limb monitoring with IMUs during standardized drills and competition (if applicable) to collect normative data in the natural sporting environment. These data will provide new opportunities to guide training and return to play programming and periodization. Also, bilateral IMUs are preferred to capture laterality differences resulting from task/position demands, limb dominance, or underlying technical deficiencies. It is also recommended that practitioners choose this approach to monitoring tactfully expressing empathy for the athletes who are already subject to a vast array of assessments that may not inform the training cycle to the degree initially intended (126).

The present review revealed a substantial lack of cohort studies, which illustrates the void in longitudinal lower limb load monitoring in sport. Observing changes throughout the competitive season poses an opportunity to garnish data that can inform training programming and periodization to avoid over- or under-stimulating athletes. Additional cohort studies may also leverage utility in examining intra-individual changes in lower limb external load and interlimb symmetry and how the competitive season, fixture congestion, and athlete fatigue influence these.

While previous investigations have noted that the shock-absorbing properties of the shoe have shown to influence tibial shock (23, 39, 40, 127), properties of playing surfaces have received similar attention (39, 42). For example, Playerload^TM^ from devices worn at the mid-tibia was significantly higher on astroturf compared to 3^rd^ generation artificial turf and natural grass among amateur soccer players performing agility drills (75). Likewise, Johnson, Outerleys (53) demonstrated that impact loads observed on a treadmill are not representative of what is seen in ecologically valid environments outdoors. It is for these reasons practitioners should seek to collect data in real-world settings.

### 4.4 Limitations

This review has several limitations that should be acknowledged. First, while the primary aim was not to focus on independent endurance athletes, given that this topic has been extensively covered in existing literature (113), this exclusion was not explicitly defined in the initial search strategy or methodology. Nevertheless, this review aligns with prior recommendations (96) advocating for the inclusion of both team and individual sports rather than limiting the scope to invasion team athletes. Second, the definition of ‘competitive’ athletes was not void of ambiguity following previously defined criteria (22). These criteria can pose challenges in referring to recreational runners as competitive athletes when they are more casually entering local races like marathons for completion. Additionally, studies relying solely on GPS or accelerometer-based data were excluded. Despite these limitations, the findings contribute to a broader understanding of athlete monitoring across different sports and competitive levels. Future research may benefit from targeting team sport athletes and wearables that have been integrated at various anatomical and implement sites.

## 5. Conclusions

The findings of this review underscore the growing adoption of IMUs in competitive sports, particularly in competitive environments, and highlight key differences in monitoring practices between categories and calibers of sport. There is no single monitoring approach that can inform practitioners on all avenues of athlete load, performance, and/or wellbeing (128). As technology continues to evolve, future research should aim to refine data interpretation, enhance ease of use, and expand longitudinal investigations to inform training. Keeping this in mind, lower limb external load may be collected in sport to enhance training, optimize rehabilitation, and create a competitive advantage via improved availability and/or optimized performance (129, 130).

## Data Availability

All relevant data are within the manuscript and its Supporting Information files.

## Acknowledgements

The authors would like to thank the Chicago Bears Football Club for their ongoing support throughout the research process.

## Supplementary Materials

None.

## Author Contributions

Author Contributions: Search phrase, inclusion and exclusion criteria, AL and JS.; literature searches, screening, and selection of articles, AL and JS; article categorization, AL; tables and figures, AL; original draft preparation, AL; review and editing, AL, JS, and TS. All authors have read and agree to the published version of the manuscript.

## Abbreviations

The following abbreviations are used in this manuscript.

IMUs: Inertial measurement units
GNSS: Global navigational satellite systems LPS Local positioning system
HAR: Human activity recognition
GRFs: Ground reaction forces
GCTs: Ground contact times

